# SLC30A8 rare variant modify contribution of common genetic and lifestyle factors toward type 2 diabetes

**DOI:** 10.1101/2024.08.28.24312708

**Authors:** Hye-Mi Jang, Mi Yeong Hwang, Yi Seul Park, Bong-Jo Kim, Young Jin Kim

**Author notes:** Corresponding authors Bong-Jo Kim; Young Jin Kim. These authors have jointly supervised this work (Bong-Jo Kim, Young Jin Kim).

## Abstract

This study aimed to investigate the modifying effects of rare genetic variants on the risk of type 2 diabetes in the context of common genetic and lifestyle factors. We conducted a comprehensive analysis of genetic and lifestyle factors associated with type 2 diabetes in a cohort of 146,284 Korean individuals. Among them, 4,603 individuals developed type 2 diabetes during the follow-up period of up to 18 years. We calculated a polygenic risk score (PRS) for type 2 diabetes and identified carriers of the rare allele I349F at SLC30A8. A Healthy Lifestyle Score (HLS) was also derived from physical activity, obesity, smoking, diet, and sodium intake levels. Using Cox proportional hazards models, we analyzed how PRS, HLS, and I349F influenced type 2 diabetes incidence. Results showed that high PRS and poor lifestyle were associated with increased risk. Remarkably, I349F carriers exhibited a lower type 2 diabetes prevalence (5.4% compared to 11.7% in non-carriers) and reduced the impact of high PRS from 23.18% to 12.70%. This trend was consistent across different HLS categories, with I349F carriers displaying a lower risk of type 2 diabetes. The integration of common and rare genetic variants with lifestyle factors enhanced type 2 diabetes predictability in the Korean population. Our findings highlight the critical role of rare genetic variants in risk assessments and suggest that standard PRS and HLS metrics alone may be inadequate for predicting type 2 diabetes risk among carriers of such variants.

**Author summary:** In our study, we investigated how rare genetic variants affect the risk of developing type 2 diabetes, particularly when combined with common genetic and lifestyle factors. We analyzed data from over 146,000 Korean individuals, following their health outcomes for up to 18 years. During this time, 4,603 participants developed type 2 diabetes. We calculated a polygenic risk score (PRS) based on common genetic variants and examined lifestyle factors such as physical activity, diet, and smoking. We also identified individuals carrying a rare genetic variant (I349F) in the SLC30A8 gene, which appeared to have a protective effect against type 2 diabetes. Our findings show that individuals with high PRS and unhealthy lifestyles are at increased risk for the disease. However, those carrying the I349F variant had a significantly lower risk, even among those with high PRS and poor lifestyle habits. This suggests that rare genetic variants can play a crucial role, and that combining genetic and lifestyle factors provides a more accurate prediction of diabetes risk. Our work highlights the importance of including rare genetic variants in personalized risk assessments for type 2 diabetes.

## Introduction

There is growing concern regarding the global burden of diabetes mellitus, which is a leading cause of mortality and morbidity (1). Type 2 diabetes, which accounts for the majority of diabetes cases, is influenced by a complex interplay between genetic and environmental factors (1). Over the past decade, genome-wide association studies (GWASs) focusing on variants with a minor allele frequency (MAF) greater than 1% have identified hundreds of loci associated with type 2 diabetes, explaining approximately half of its known heritability (2–4). Polygenic risk scores (PRS) based on these associations have been used to summarize individual genetic risk by considering the number and effect sizes of risk alleles. Individuals in genetically high-risk groups showed approximately 2–3 times higher prevalence of type 2 diabetes compared to those in the remaining groups (2–6). However, a recent study has highlighted the significant impact of rare variants (MAF < 1%) with substantial genetic effects, resulting in a nearly 50% reduction in the prevalence of type 2 diabetes among individuals carrying the rare allele (7). These studies are expected to contribute to the comprehensive identification of high-risk groups for type 2 diabetes and implementation of appropriate interventions (6, 7).

In addition to the aforementioned genomic efforts, numerous studies have identified environmental factors associated with type 2 diabetes (8). Recent investigations have proposed healthy lifestyle score (HLS), which integrates individual risk factors, such as physical activity, dietary habits, smoking status, and alcohol consumption, as a means of assessing an individual’s risk of developing type 2 diabetes (9–12). Individuals with an “unfavorable” lifestyle showed an increased incident type 2 diabetes compared with the baseline group (10–12). A risk-stratified subset of individuals based on PRS and HLS showed varied levels of incident type 2 diabetes, suggesting an additive contribution of genetic and environmental factors to the susceptibility (10–12). However, despite the marked impact of rare variants on type 2 diabetes, previous studies have primarily focused on PRS derived from common variants and their interactions with HLS (7, 9–12). Currently, the integration of PRS, HLS, and rare allele for type 2 diabetes risk stratification is limited because of insufficient large-scale genomic information with comprehensive coverage of rare variants across the human genome.

To explore the potential combined impact of PRS, HLS, and rare variants, we analyzed 146,284 samples from the Korean Genome and Epidemiology Study (KoGES) genotyped using the Korea Biobank Array (KBA). Previous GWASs utilizing KBA have demonstrated varying levels of type 2 diabetes prevalence for different combinations of common and rare genetic factors (7). In this study, we have performed association analyses for common and rare variants associated with type 2 diabetes and developed models that incorporated PRS, HLS, and discovered rare variant. Our study highlights that the rare allele modify the contributions of common genetic and lifestyle factors to type 2 diabetes and emphasizes that enhanced type 2 diabetes risk stratification can be achieved by integrating PRS, HLS, and a rare allele.

## Results

### GWAS on type 2 diabetes in the Korean population

The demographic characteristics of 123,822 samples analyzed in this study are summarized in **Supplementary Table 1**. Logistic regression analysis was conducted to identify the variants associated with type 2 diabetes in the Korean population. Among the 8.3 M high-quality imputed common variants (MAF >= 1%), 44 independent loci were identified with a *p*-value < 5×10^-8^ (**Supplementary Figure 2 & Supplementary Table 3**). These loci were found within a 1-Mb window of previously known loci (**supplementary Table 3**) ((3),(13)).

To investigate the contribution of rare variants to type 2 diabetes, logistic regression analysis was performed at the single-variant level, and a burden test using SKAT-O was conducted at the gene level (14). Only the I349F variant in *SLC30A8*, previously discovered in the Korean population, showed an association with T2D (OR=0.40, *p*=7.30×10^-16^) with a *p*-value < 5×10^-8^ as a threshold (**Supplementary Table 4)**. A gene-based test identified four gene associations (*p* < 1×10^-5^) involving *PSMB8*, *PSMB8-AS1*, *SLC30A8*, and *TAP1* (**Supplementary Table 4**). Among the variants used in the gene-based test, four, including I349F, were marginally significant (*p* < 0.05) **(Supplementary Table 5)**. Previously, some rare variants have shown non-independent associations owing to the correlated genetic architecture of nearby common and rare signals (7). After adjusting for nearby common signals (rs56118007 at *MHC* and rs13266634 at *SLC30A8*), only the association with *SLC30A8* remained significant **(Supplementary Table 4)**.

To replicate the rare associations identified in the discovery dataset containing 124 K Korean samples, an independent set of 22,462 samples was genotyped to validate the results of single-variant and burden tests. In the replication dataset, I349F showed a significant association (OR=0.55, *p*=1.73×10^-2^) with consistent directionality of effect size (**Supplementary Table 4**). However, the gene-based test in the replication dataset did not yield any significant association, possibly owing to the small number of rare carriers (**Supplementary Table 4**). Based on the results of the rare variant analysis (**Supplementary Tables 4 & 5**), a rare variant, I349F, with a protective effect against type 2 diabetes was selected in *SLC30A8* to subset the group of rare-allele carriers. Among the 146,284 individuals, rare-allele carriers had a type 2 diabetes prevalence of 5.7%, whereas non-carriers had a prevalence of 11.7% (**Supplementary Table 6**). For incident T2D cases, rare-allele carriers had a type 2 diabetes incidence of 4.7% and 7.3% for carriers and non-carriers, respectively (**Supplementary Table 6**).

### Polygenic prediction of type 2 diabetes and its modification by rare allele

PRS were calculated for KoGES participants using summary statistics from an independently conducted GWAS for type 2 diabetes at Biobank Japan (15). The constructed type 2 diabetes-PRS showed a strong association with prevalent type 2 diabetes cases in 146,284 individuals of the KoGES (OR=2.06, P=1.65×10^-1017^), explaining 8.81% of the variance (**Table 1**). Type 2 diabetes prevalence increased as the type 2 diabetes-PRS increased (**Supplementary Figure 3**). When comparing the group of individuals with the top 1% type 2 diabetes-PRS to the median group (40– 60%), the genetically high-risk group showed an approximately a six-fold increase in type 2 diabetes prevalence (**Table 1**). The top 5% and 10% conferred a 4.2- and 3.7-fold increase in type 2 diabetes prevalence, respectively. Among the all individuals without type 2 diabetes at the baseline, type 2 diabetes-PRS showed a strong association with incident type 2 diabetes cases (HR=1.52, P=1.49x10^-168^) (**Table 1**). Similar to those of prevalent type 2 diabetes cases, the top 1% and 5% conferred 2.6- and 2.3-fold increase in type 2 diabetes incidence, respectively (**Table 1**).

**Table 1.** Impact of high T2D-PRS in the Korean population.

| PRS |  | Prevalent T2D |  |  |  |  |  | Incident T2D |  |  |  |
| --- | --- | --- | --- | --- | --- | --- | --- | --- | --- | --- | --- |
|  |  | Case (N) | Control (N) | Odds ratio | P-value | 95% CI | Pseudo R<sup>2</sup> | Case (N) | Hazard ratio | P-value | 95% CI |
| T2D-PRS |  | 13,220 | 100,453 | 2.06 | 1.65E-1017 | 2.01-2.10 | 8.81% | 4,603 | 1.52 | 1.49E-168 | 1.48 - 1.57 |
| High Risk Group | Reference Group | Case (N) | Control (N) | Odds ratio | P-value | 95% CI | Pseudo R<sup>2</sup> | Case (N) | Hazard ratio | P-value | 95% CI |
| Top 20 % | Remaining 80% | 4,920 | 16,407 | 3.34 | 1.27E-719 | 3.21-3.48 | 5.53% | 1,300 | 1.97 | 2.42E-95 | 1.85-2.10 |
| Top 10% | Remaining 90% | 2,849 | 7,645 | 3.72 | 2.13E-568 | 3.53-3.91 | 4.15% | 691 | 2.10 | 3.65E-72 | 1.94-2.28 |
| Top 5% | Remaining 95% | 1,568 | 3,545 | 4.19 | 3.63E-392 | 3.93-4.48 | 2.78% | 375 | 2.31 | 2.94E-54 | 2.08-2.57 |
| Top 1% | Remaining 99% | 399 | 595 | 6.09 | 1.12E-148 | 5.32-6.98 | 1.04% | 73 | 2.61 | 4.70E-16 | 2.07-3.29 |

In a previous study, we demonstrated that the I349F variant at *SLC30A8* modified the effect of the common variant–based genetic risk score, resulting in a decrease in type 2 diabetes prevalence by approximately half, regardless of the score (7). As expected, the modification effect of the rare allele was shown to be additive to that of the common variant–based PRS (**Figure 1**). For example, among the samples in the discovery study, the type 2 diabetes prevalence of in the top quintile of the type 2 diabetes-PRS decreased from 23.2% overall to 12.7% in individuals carrying rare protective allele of *SLC30A8*, and from 4.6% to 1.8% in the bottom quintile (**Supplementary Table 6**).

**Figure 1.**
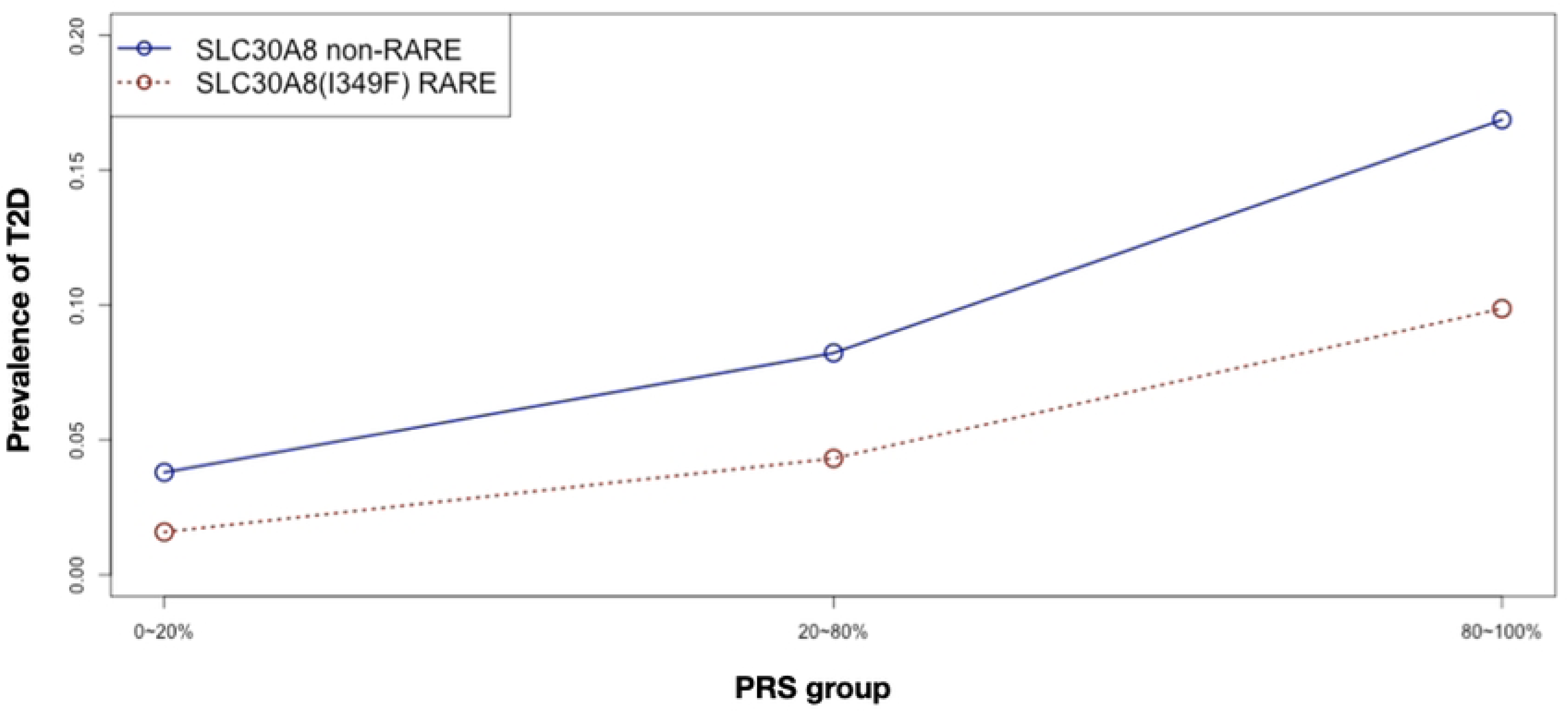
Modified effect of type 2 diabetes-PRS by protective rare allele. After sorting the type 2 diabetes-PRS scores in increasing order, the PRS bins were categorized as 1^st^ bin (1–20%), 2^nd^ bin (21–80%), and 3^rd^ bin (81–100%). For rare-allele carriers and non-carriers, all samples were categorized into three type 2 diabetes-PRS bins, and the prevalence of type 2 diabetes was calculated separately for rare-allele carriers and non-carriers.

The type 2 diabetes-PRS and subsets of rare allele were examined further to assess their ability to predict future type 2 diabetes. As shown in **Supplementary Table 7**, participants with intermediate and high type 2 diabetes-PRS had a hazard ratio (HR) of 1.83 (*p*=4.30×10^-38^) and 3.16 (*p*=7.86×10^-113^) for incident type 2 diabetes, respectively, compared to individuals in the bottom 20%. When considering the presence of the protective rare allele in addition to type 2 diabetes-PRS, non-carriers with intermediate and high PRS had an HR of 1.84 (P=7.07×10^-39^) and 3.17 (P=1.09×10^-112^) for incident type 2 diabetes, respectively, compared to the bottom 20%.

Meanwhile, carriers with intermediate and high PRS had an HR of 0.84 (*p*=5.01×10^-1^) or 2.60 (*p*=2.59×10^-4^) for incident type 2 diabetes, respectively, compared to individuals in the bottom 20% of type 2 diabetes-PRS.

### Contribution of a healthy lifestyle to type 2 diabetes

Among the five lifestyle-related factors, 146,284 individuals who followed a healthy lifestyle constituted 86.32%, 65.88%, 35.71%, 45.34%, and 39.65% in terms of current smoking status, obesity, healthy diet, physical activity, and sodium-intake status, respectively (**Supplementary Table 8**). All five healthy-lifestyle factors showed protective effects against incident type 2 diabetes and four factors except sodium intake were statistically significant (*p* < 0.05) (**Supplementary Table 8**). These results suggest that these lifestyle factors could be adequate predictors of future type 2 diabetes.

Based on the calculated HLS, 35,743 (24.43%), 91,607 (62.62%), and 18,934 (12.94%) individuals were in the favorable, intermediate, and unfavorable groups, respectively (**Table 2**). The HLS-unfavorable group showed an approximately 2.5-fold increase in incident type 2 diabetes compared to the favorable group (HR=2.48, *p*=6.31×10^-51^) (**Table 2**). The HLS-intermediate group showed a less pronounced increase in incident type 2 diabetes (HR=1.45, *p*=8.04×10^-16^).

**Table 2.** Impact of high T2D-PRS in the Korean population.

| Group | Lifestyle score | Total (N (%)) | Prevalent T2D |  |  |  | Incident T2D |  |  |  |
| --- | --- | --- | --- | --- | --- | --- | --- | --- | --- | --- |
|  |  |  | Case (N) | Odds ratio | P-value | 95% CI | Case (N) | Hazard ratio | P-value | 95% CI |
| **Favorable (baseline)** | 4,5 | 35,743 (24.43) | 2,682 | - | - | - | 620 | - | - | - |
| **Intermediate** | 2,3 | 91,607 (62.62) | 8,263 | 1.19 | 2.65E-12 | 1.13-1.25 | 2,818 | 1.45 | 8.04E-16 | 1.32-1.58 |
| **Unfavorable** | 0,1 | 18,934 (12.94) | 2,275 | 1.67 | 3.87E-46 | 1.55-1.79 | 1,165 | 2.48 | 6.31E-51 | 2.20-2.79 |

### Combinatorial effect of genetic factors and HLS on future type 2 diabetes

To assess the combined effect of PRS, rare allele, and HLS, we stratified all 146,284 individuals into groups considering three PRS groups, presence of rare allele, and three HLS groups. Unfortunately, the combination of all three factors showed less than five counts for groups of rare-allele carriers with an unfavorable lifestyle and high PRS. Therefore, we focused only on the combinatorial effects of the PRS-HLS and HLS-rare allele on future type 2 diabetes.

Compared with the reference (low PRS and favorable HLS), among 130,590 individuals without type 2 diabetes at the baseline, incident type 2 diabetes risk was increased as the risk of either PRS or HLS increased (**Figure 2**, **Table 3**). For example, high HLS showed increased type 2 diabetes risk over the intermediate group, regardless of the PRS group. PRS and HLS showed an increasing tendency toward type 2 diabetes risk in a roughly additive manner. Moreover, individuals with an unfavorable lifestyle and the top 20% of type 2 diabetes-PRS had a highest risk of incident type 2 diabetes (HR=9.48, *p*=1.39×10^-43^) (**Table 3**). Similar patterns were also observed for prevalent type 2 diabetes **(Supplementary Figure 4)**. These results were consistent with those of previously conducted studies (10, 16).

**Figure 2.**
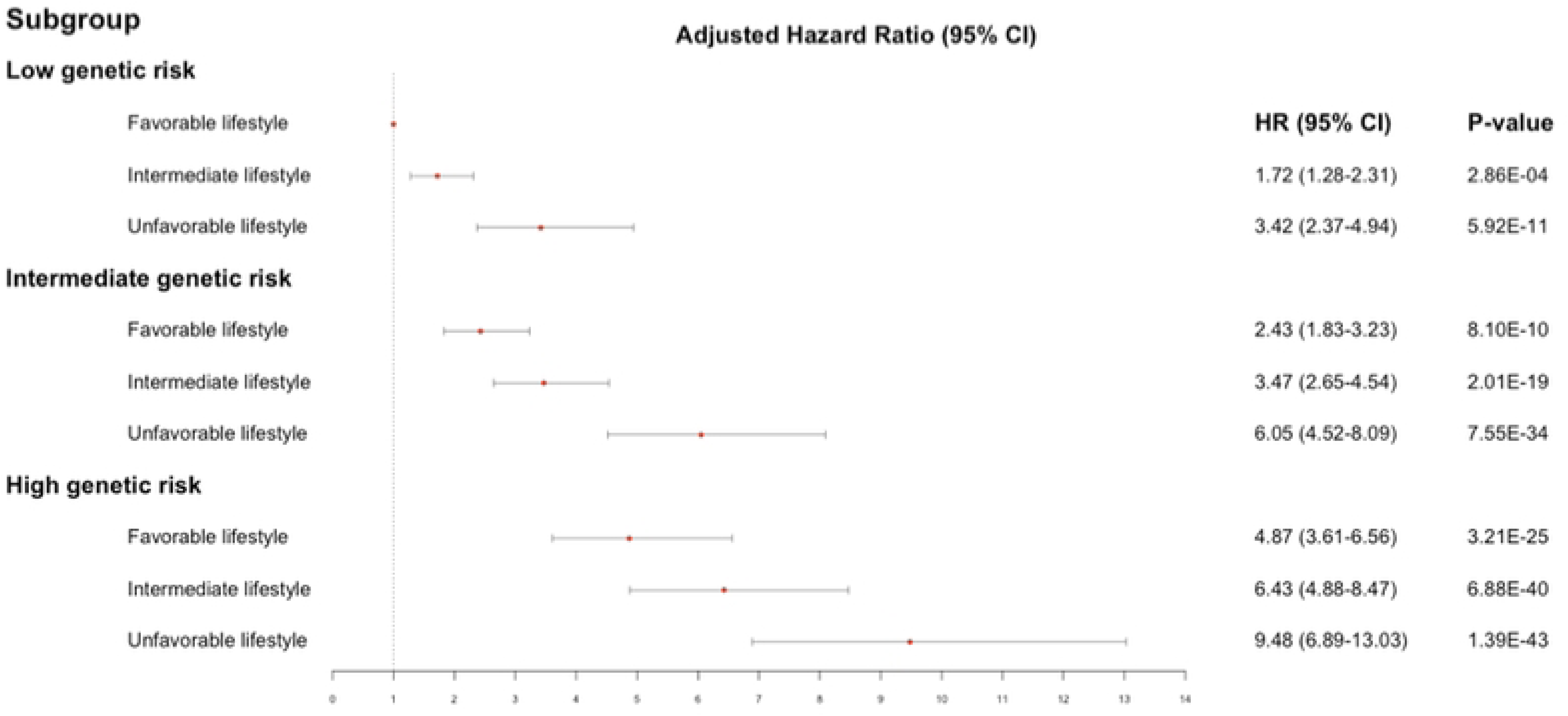
Risk of future type 2 diabetes mellitus according to genetic and lifestyle risk factors. Associations among type 2 diabetes-PRS, HLS, and incident type 2 diabetes. The analyses were adjusted for age, sex, and the area of recruitment.

**Table 3.** Predictability of T2D-PRS and HLS for future T2D.

| PRS group | HLS group | Cases (N) | Hazard ratio | P-value | 95% CI |
| --- | --- | --- | --- | --- | --- |
| Bottom 20% | Favorable | 55 | - | - | - |
|  | Intermediate | 345 | 1.72 | 2.86E-04 | 1.28-2.31 |
|  | Unfavorable | 151 | 3.42 | 5.92E-11 | 2.37-4.94 |
| 20-80% | Favorable | 364 | 2.43 | 8.10E-10 | 1.83-3.23 |
|  | Intermediate | 1666 | 3.47 | 2.01E-19 | 2.65-4.54 |
|  | Unfavorable | 722 | 6.05 | 7.55E-34 | 4.52-8.09 |
| Top 20% | Favorable | 201 | 4.87 | 3.21E-25 | 3.61-6.56 |
|  | Intermediate | 807 | 6.43 | 6.88E-40 | 4.88-8.47 |
|  | Unfavorable | 292 | 9.48 | 1.39E-43 | 6.89-13.03 |

Each group was compared with the baseline (Favorable HLS and Bottom 20% T2D-PRS)

Studying the combinatorial effect of HLS and presence of rare allele was limited owing to the small number of incident type 2 diabetes cases in the stratified groups. By using the baseline group with favorable HLS and non-carriers, non-carriers and carriers among the unfavorable and intermediate-HLS groups had an HR of 1.59 (*p*=7.88×10^-25^) and 0.84 (*p*=3.97×10^-1^), respectively, for incident type 2 diabetes (**Table 4**). Although the results from the group with rare allele carriers were not statistically significant, protective rare-allele carriers showed a decreased risk of incident type 2 diabetes compared with the baseline group. **Figure 3** shows that individuals carrying rare protective allele within the unfavorable or intermediate-HLS group exhibited a similar trend in survival rates compared with non-carriers within the favorable-HLS group. When these analytical schemes were applied to prevalent type 2 diabetes cases, the combination of HLS and presence of rare allele showed patterns similar to those of the HLS and PRS relationship by acting additively with each other (*p* < 1.20×10^-6^, **Table 4**). Compared to the baseline (favorable HLS and non-carriers), rare-allele carriers showed a decreased type 2 diabetes risk compared to non-carriers, regardless of the HLS group (**Supplementary Table 9**). For instance, non-carriers and carriers with intermediate HLS had an OR of 1.18 (*p*=8.03×10^-12^) and 0.47 (*p*=6.28×10^-7^) for prevalent type 2 diabetes, respectively, compared to the baseline group (**Table 4**).

**Figure 3.**
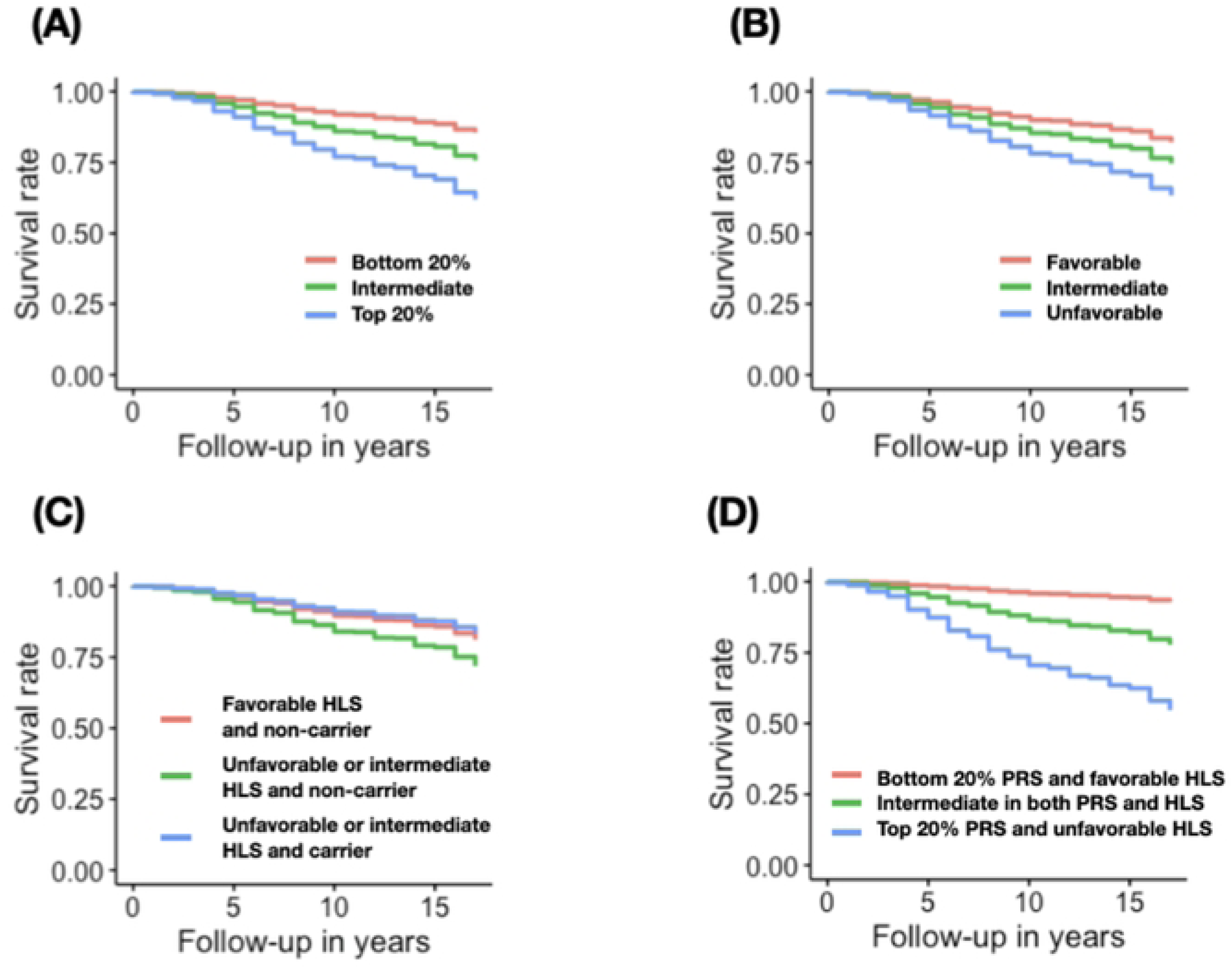
Survival rate of incident type 2 diabetes mellitus according to genetic and lifestyle risk factors. Survival rate of incident type 2 diabetes mellitus, stratified by (A) Type 2 diabetes-PRS (bottom 20%, intermediate 20–80%, and top 20%), (B) HLS (favorable, intermediate, and unfavorable), (C) HLSxRare allele combination (favorable HLS & non-carrier, unfavorable or Intermediate HLS & non-carrier, and unfavorable or Intermediate HLS & carrier), and (D) Type 2 diabetes-PRSxHLS combination (bottom 20% PRS & favorable HLS, intermediate level in both PRS and HLS, and top 20% PRS & unfavorable).

**Table 4.** Predictability of T2D-PRS and HLS for future T2D.

| HLS group | I349F | Prevalent T2D |  |  |  | Incident T2D |  |  |  |
| --- | --- | --- | --- | --- | --- | --- | --- | --- | --- |
|  |  | Case (N) | Odds ratio | P-value | 95% CI | Case (N) | Hazard ratio | P-value | 95% CI |
| Favorable | non-carrier | 2,659 | - | - | - | 613 | - | - | - |
| Intermediate + Unfavorable | non-carrier | 10,422 | 1.25 | 1.22E-20 | 1.19-1.31 | 3,947 | 1.59 | 7.88.E-25 | 1.45-1.74 |
|  | Carrier | 69 | 0.53 | 1.02E-06 | 0.41-0.68 | 27 | 0.84 | 3.97.E-01 | 0.57-1.25 |

Each group was compared to the baseline (Favorable HLS & non-carriers)

When the linear model was constructed based on HLS, PRS, and rare allele, the combination of these factors showed increased predictability for incident type 2 diabetes over the baseline model that consisted of age and sex. The C-index analysis provided increased predictability over the baseline model with C-index values of 0.67, 0.64, 0.62, and 0.69 for PRS, HLS, rare allele, and combinations, respectively, while the baseline model showed an C-index value of 0.59. These results demonstrated the importance of various factors in explaining different aspects of type 2 diabetes.

## Discussion

Our study offers valuable insights into the relationship between genetic and lifestyle factors and risk of type 2 diabetes in the Korean population. We identified significant genetic variants, including common and rare allele, that contribute to this risk. Additionally, we demonstrated the crucial role of a healthy lifestyle in reducing the incidence of type 2 diabetes. Moreover, we highlighted the modified effects of rare protective allele on genetic and lifestyle factors related to type 2 diabetes, emphasizing the importance of considering various aspects of genetics and lifestyle factors in predictive models.

To the best of our knowledge, this is the first study to introduce the modification effect of rare variant on common genetic and lifestyle factors related to incident type 2 diabetes. We showed that rare variant have a substantial impact on type 2 diabetes in carriers of the rare allele. Furthermore, the effects of HLS were also modified by these rare protective allele. Despite the recent large-scale sequencing studies by Cao et al. and Halldorsson et al. (17, 18) have demonstrated that more than 94% of variants in the human genome are rare (MAF < 1%), studies regarding rare variants are still in their early stages owing to the limited availability of large-scale sequencing data. Considering the vast number of rare variants in the human genome, studying these variants is crucial for accurately assessing the risk of various diseases. Additionally, because most rare variants are likely to be specific to particular populations (7, 17, 18), further sequencing efforts in diverse populations are needed to identify rare variants with strong modification effects on various risk factors and type 2 diabetes.

Herein, we have provided insights into the risk stratification of type 2 diabetes and offered clues for personalized treatment based on individual genetics and lifestyles. **Figure 3** illustrates the survival rate of incident type 2 diabetes based on Cox regression models for risk factors analyzed in this study. Overall, individuals who do not carry rare protective allele, have an unfavorable lifestyle, and have high type 2 diabetes-PRS may develop type 2 diabetes at an early age. Based on an individual’s specific risk profile, personalized treatment is possible by implementing appropriate lifestyle interventions for individuals with unfavorable lifestyles but with a genetic risk lower than the top 80% of type 2 diabetes-PRS. Conversely, routine screening of individuals with high genetic risk should be conducted regardless of their lifestyle. Furthermore, the stratification strategy employed in this study complements recent efforts in subtyping type 2 diabetes. A more accurate subclassification of type 2 diabetes could provide a solid framework for future research on its pathogenesis and personalized medicine for diabetes, as demonstrated by Ahlqvist et al. (19).

We acknowledge that there are some limitations to this study. First, the sample size for the replication analysis of rare variants and assessment of combinatorial effects was relatively small, which may have limited the statistical power to detect significant associations. Second, the lifestyle factors assessed in our study were self-reported and are subject to potential bias. Future studies with larger sample sizes and a more comprehensive assessment of lifestyle factors would further enhance our understanding of the complex interplay between genetics, lifestyle, and the risk of type 2 diabetes. Finally, considering the population-specific nature of rare variants, the results of this study may not be directly applicable to other populations. The rare variant discovered at *SLC30A8* were found to be polymorphic in East Asians, yet monomorphic in other populations.

In conclusion, our findings highlight the importance of genetic variants, including common and rare alleles, in the development of type 2 diabetes in Korean population. The combination of common and rare genetic factors along with lifestyle factors improves the predictability of type 2 diabetes. These results contribute to our understanding of the etiology of type 2 diabetes and may have implications for personalized prevention and intervention strategies for individuals at risk of developing this disease. By considering the interplay between genetics and lifestyle, we can better identify individuals who may benefit from targeted interventions and tailored treatment options.

### Materials and Methods Study subjects

This study was approved by the institutional review board of the Korea Disease Control and Prevention Agency, Republic of Korea. In the Korean Genome and Epidemiology Study (KoGES), 211,725 participants were recruited from three population-based cohorts: the KoGES_Ansan and Ansung study (n=10,030), KoGES_ Health Examinee study (HEXA, n=173,357), and KoGES_Cardiovascular Disease Association Study (CAVAS, n=28,338). KoGES has been described previously (20, 21). Numerous variables, including epidemiological surveys, physical examinations, and laboratory tests, were examined. All participants aged 40–70 years provided written informed consent.

The type 2 diabetes cases and controls were defined according to the American Diabetes Association criteria. The cases included those with a fasting plasma glucose (FPG) concentration ≥ 126 mg/dL (7.0 mmol/L), an oral glucose tolerance test (OGTT) ≥ 200 mg/dL (11.1 mmol/L), or an HbA1c ≥ 6.5% (48 mmol/mol). A participant who reported type 2 diabetes treatment was included as a case. Controls included subjects without a history of diabetes who satisfied the following criteria: FPG concentration < 100 mg/dL (5.6 mmol/L), OGTT < 140 mg/dL (7.8 mmol/L), and HbA1c level < 6% (42 mmol/mol). The variables of OGTT and HbA1c were used, if available. Among the genotyped subjects in the discovery study (n=123,822), 11,087 cases and 86,058 controls were selected for further analyses (**supplementary Table 1**).

Among the all genotyped participants (n=146,284), incident cases of type 2 diabetes were identified from those who did not have it at baseline recruitment (**Supplementary Table 1**).

Among 130,590 participants without type 2 diabetes at baseline, 4,603 samples were regarded as incident cases if they met one of the following criteria during the 18 years of follow-up: past diagnosis or prior type 2 diabetes treatment, FPG ≥ 126 mg/dL, OGTT ≥ 200 mg/dL, or HbA1c ≥ 6.5%.

### Genotyping and quality control

Among KoGES participants, 134,721 samples were genotyped using the KBA, an optimized single-nucleotide polymorphism (SNP) microarray for genome studies in the Korean population (21). Details of genotyping and quality control have been described previously (7). Briefly, 123,822 samples, with informed consent at the time of the analysis and non-missing phenotypes, were retained for further analysis after quality-control processes based on the following criteria for each batch grouped by versions (v1.0 and v1.1) of KBA: [1] samples were excluded if gender discrepancy, low call rate (<97%), excessive heterozygosity, 2^nd^-degree related samples, and outliers of principle component analysis 2) variants were excluded for low call rate (<95%), Hardy Weinberg equilibrium (HWE) failure (*p* < 10^-6^), and low minor allele frequency (MAF) (< 1%). Therefore, less than 550 K SNPs were retained for the phasing and imputation analyses. To study rare genotyped variants, 68,431 rare functional autosomal variants (MAF < 1%) were used for further analysis after quality control. Among approximately 160 K initial functional variants (missense, frameshift, start/stop gain or loss, splice site donor or acceptor, and structural interaction), rare variants were filtered out for allele frequency discrepancy [> 0.5% either one of all batches, 2,579 sequenced Korean samples (7), 504 East Asian samples from the 1,000 Genomes Project Phase 3 (22), and 9,435 East Asian samples from the gnomAD database (23) ], minor allele count < 30, HWE failure (*p* < 10^-6^), and missing rate (>30%).

### Genotype imputation

A pre-phasing-based imputation analysis was performed on the QCed data. Eagle v2.3 (24) was used to phase the QCed data, and the phased data were imputed using Impute v4 (25) with a merged reference panel of 2,504 samples from the 1,000 Genomes Phase 3 (22) and 397 samples from the Korean Reference Genome (21). The GEN-formatted file, an output from Impute v4, was converted to the VCF format with imputed dosages using GEN2VCF (26). For further analysis, 8.3 M high-quality imputed common variants were obtained by excluding variants with imputation quality < 0.8 and MAF < 1%.

### Replication study

For the replication study, approximately 24,000 samples from the HEXA cohort, a part of KoGES, were genotyped using the KBA, and 22,462 samples, with informed consent at the time of the analysis and non-missing phenotypes, were retained after quality-control procedures described above (7). For further analysis, 8.1 M high-quality imputed variants were used after phasing and imputation analysis. The HLS for the replication dataset was calculated using the aforementioned protocol.

### Calculation of PRS

PRS was calculated for all individuals analyzed in this study (n=146,284). To construct the PRS for type 2 diabetes, adjusted weights were obtained using PRS-CS (27) with type 2 diabetes GWAS conducted by Biobank Japan (15). About 970 K HapMap phase 3 variants were used to calculate the PRS using adjusted weights. The calculated PRS values were transformed to follow a normal distribution. Based on the PRS, individuals were categorized into three groups: low (bottom 20%), intermediate (20–80%), and high (80–100%).

### Construction of healthy lifestyle score

The healthy lifestyle score was calculated for all individuals analyzed in this study (n=146,284). To measure the magnitude of the healthy lifestyle of a participant, five healthy lifestyle–related factors were assessed considering those from previous literature (9, 11, 28) and the disease burden owing to high sodium intake in Korea (28, 29). These lifestyle factors were physical activity, obesity, smoking status, healthy dietary patterns, and sodium intake status as retrieved from the survey questionnaires. Each factor was coded as 1 (healthy lifestyle) if it met the criteria and 0 (unhealthy lifestyle) otherwise. The HLS for each participant was calculated by summing all five factors. An individual was regarded as having healthy lifestyle based on the following criteria: 1) physical activity at least 30 min once a week, 2) body mass index < 25 kg/m^2^, 3) current non-smoker, 4) healthy dietary pattern (reaching the daily recommended in ≥ 5 out of 9 items **(supplementary Table 2)**, and 5) daily sodium intake < 2 g. Finally, HLS ranging from 0 to 5 were categorized into three groups: unfavorable (HLS = 0–1), intermediate (HLS = 2–3), and favorable (HLS = 4–5).

### Statistical analysis

Single-variant association analysis (logistic regression) for type 2 diabetes was performed using Hail v0.2.126, assuming an additive mode of inheritance based on alternative allele count, adjusting for age, sex, and recruitment area. Cluster plots of the associated rare variants from single variants and gene-based test results were visually inspected (*SLC30A8* variants in **Supplementary Figure 1).** An inverse variance–weighted meta-analysis was performed using METAL software (30) by combining datasets of KBAv1.0 and KBAv1.1. A locus was defined by clustering variants (*p*< 5×10^-8^) within a 500-kb range. Gene-based burden test for rare functional variants was performed using the optimal unified test (SKAT-O) (14). A lead signal of the locus was selected as the most significant variant within the locus of clustered associated variants (*p*≤ 5×10^-8^) if the variants were located within a 500-kb range. A logistic regression model was used to test the association between genetic and prevalent type 2 diabetes cases adjusting for age, sex, and recruitment area. A Cox proportional hazards model using the R package “survival” was used to test the association between genetic or lifestyle factors and incident type 2 diabetes events adjusting for age, sex, and recruitment area (31)

### Data and resource availability

The summary-level results generated in this study are available on the KNIH (Korea National Institute of Health) PheWeb website (https://coda.nih.go.kr/usab/pheweb/intro.do).

## Acknowledgements

The KBA data were provided by the Collaborative Genome Program for Fostering New Post-Genome Industry (3000-3031b).

## Author Contributions

B.-J.K. and Y.J.K. conceived and designed the study. H.-M.J., B.-J.K., and Y.J.K. wrote the manuscript. H.-M.J., M.Y.H., Y.S.P., and Y.J.K. analyzed the data. All the authors have interpreted the results, edited the manuscript, and approved the submission of the final version of the article for publication.

## Funding

This study was supported by intramural grants from the National Institute of Health, Republic of Korea (grant numbers 2019-NI-097-02, 2022-NI-065-01, and 2022-NI-067-01).

## Conflicts of Interest

No potential conflicts of interest relevant to this article were reported.

## Supporting information captions

### Supplementary Tables

**Supplementary table 1. Demographic characteristics of study samples**

**Supplementary table 2. Healthy dietary items**

**Supplementary table 3. Common variants associated with Type 2 diabetes in the discovery stage**

**Supplementary table 4. Rare variant association results**

**Supplementary table 5. Single variant associations (genes from burden test)**

**Supplementary table 6. T2D-PRS and its modification by protective rare allele**

**Supplementary table 7. Predictability of T2D-PRS and rare allele for future T2D**

**Supplementary table 8. Impact of individual lifestyle factors on T2D**

**Supplementary table 9. Combinatorial effect of HLS and rare allele on prevalent T2D**

### Supplementary Figures

**Supplementary Figure 1. Cluster plots of rare variant at *SLC30A8***

(A) 8:118184855_A/T (I349F) discovery study, (B) 8:118184855_A/T (I349F) replication study

**Supplementary Figure 2. Manhattan plot of type 2 diabetes GWAS**

Manhattan plot shows logistic regression analysis results of common variants for T2D. Red horizontal line indicates -log10(5e-8). Blue and orange colors indicate difference chromosomes.

**Supplementary Figure 3. Prevalence of type 2 diabetes by T2D-PRS group**

Samples were grouped into 30 groups based on PRS scores in an increasing order. For each PRS bin, T2D prevalence was calcuated as # of T2D samples divided by # of samples in the PRS bin.

**Supplementary Figure 4. Risk of prevalent type 2 diabetes according to genetic and lifestyle risk factors**

Association of T2D-PRS and HLS with prevalent T2D. Analyses were adjusted for age, sex, and recruitment area.

## Notes

### Competing Interest Statement

The authors have declared no competing interest.

### Author Declarations

This study was approved by the institutional review board of the Korea Disease Control and Prevention Agency, Republic of Korea.

## References

1. Lin X, Xu Y, Pan X, Xu J, Ding Y, Sun X, et al. Global, regional, and national burden and trend of diabetes in 195 countries and territories: an analysis from 1990 to 2025. Sci Rep. 2020;10(1):14790.

2. Mahajan A, Taliun D, Thurner M, Robertson NR, Torres JM, Rayner NW, et al. Fine-mapping type 2 diabetes loci to single-variant resolution using high-density imputation and islet-specific epigenome maps. Nat Genet. 2018;50(11):1505–13.

3. Spracklen CN, Horikoshi M, Kim YJ, Lin K, Bragg F, Moon S, et al. Identification of type 2 diabetes loci in 433,540 East Asian individuals. Nature. 2020;582(7811):240-5.

4. Mahajan A, Spracklen CN, Zhang W, Ng MCY, Petty LE, Kitajima H, et al. Multi-ancestry genetic study of type 2 diabetes highlights the power of diverse populations for discovery and translation. Nat Genet. 2022;54(5):560–72.

5. Khera AV, Chaffin M, Aragam KG, Haas ME, Roselli C, Choi SH, et al. Genome-wide polygenic scores for common diseases identify individuals with risk equivalent to monogenic mutations. Nat Genet. 2018;50(9):1219–24.

6. Torkamani A, Wineinger NE, Topol EJ. The personal and clinical utility of polygenic risk scores. Nat Rev Genet. 2018;19(9):581–90.

7. Kim YJ, Moon S, Hwang MY, Han S, Jang HM, Kong J, et al. The contribution of common and rare genetic variants to variation in metabolic traits in 288,137 East Asians. Nat Commun. 2022;13(1):6642.

8. Ismail L, Materwala H, Al Kaabi J. Association of risk factors with type 2 diabetes: A systematic review. Comput Struct Biotechnol J. 2021;19:1759–85.

9. Khera AV, Emdin CA, Drake I, Natarajan P, Bick AG, Cook NR, et al. Genetic Risk, Adherence to a Healthy Lifestyle, and Coronary Disease. N Engl J Med. 2016;375(24):2349–58.

10. Said MA, Verweij N, van der Harst P. Associations of Combined Genetic and Lifestyle Risks With Incident Cardiovascular Disease and Diabetes in the UK Biobank Study. JAMA Cardiol. 2018;3(8):693–702.

11. Schnurr TM, Jakupovic H, Carrasquilla GD, Angquist L, Grarup N, Sorensen TIA, et al. Obesity, unfavourable lifestyle and genetic risk of type 2 diabetes: a case-cohort study. Diabetologia. 2020;63(7):1324–32.

12. Li H, Khor CC, Fan J, Lv J, Yu C, Guo Y, et al. Genetic risk, adherence to a healthy lifestyle, and type 2 diabetes risk among 550,000 Chinese adults: results from 2 independent Asian cohorts. Am J Clin Nutr. 2020;111(3):698–707.

13. Vujkovic M, Keaton JM, Lynch JA, Miller DR, Zhou J, Tcheandjieu C, et al. Discovery of 318 new risk loci for type 2 diabetes and related vascular outcomes among 1.4 million participants in a multi-ancestry meta-analysis. Nat Genet. 2020;52(7):680–91.

14. Lee S, Emond MJ, Bamshad MJ, Barnes KC, Rieder MJ, Nickerson DA, et al. Optimal unified approach for rare-variant association testing with application to small-sample case-control whole-exome sequencing studies. Am J Hum Genet. 2012;91(2):224–37.

15. Suzuki K, Akiyama M, Ishigaki K, Kanai M, Hosoe J, Shojima N, et al. Identification of 28 new susceptibility loci for type 2 diabetes in the Japanese population. Nature Genetics. 2019;51(3):379–86.

16. Langenberg C, Sharp SJ, Franks PW, Scott RA, Deloukas P, Forouhi NG, et al. Gene-lifestyle interaction and type 2 diabetes: the EPIC interact case-cohort study. PLoS Med. 2014;11(5):e1001647.

17. Cao Y, Li L, Xu M, Feng Z, Sun X, Lu J, et al. The ChinaMAP analytics of deep whole genome sequences in 10,588 individuals. Cell Res. 2020;30(9):717–31.

18. Halldorsson BV, Eggertsson HP, Moore KHS, Hauswedell H, Eiriksson O, Ulfarsson MO, et al. The sequences of 150,119 genomes in the UK Biobank. Nature. 2022;607(7920):732-40.

19. Ahlqvist E, Prasad RB, Groop L. Subtypes of Type 2 Diabetes Determined From Clinical Parameters. Diabetes. 2020;69(10):2086–93.

20. Kim Y, Han B-G, the Ko GESg. Cohort Profile: The Korean Genome and Epidemiology Study (KoGES) Consortium. International Journal of Epidemiology. 2017;46(2):e20-e.

21. Moon S, Kim YJ, Han S, Hwang MY, Shin DM, Park MY, et al. The Korea Biobank Array: Design and Identification of Coding Variants Associated with Blood Biochemical Traits. Scientific Reports. 2019;9(1):1382.

22. Auton A, Abecasis GR, Altshuler DM, Durbin RM, Abecasis GR, Bentley DR, et al. A global reference for human genetic variation. Nature. 2015;526(7571):68-74.

23. Karczewski KJ, Francioli LC, Tiao G, Cummings BB, Alföldi J, Wang Q, et al. The mutational constraint spectrum quantified from variation in 141,456 humans. Nature. 2020;581(7809):434-43.

24. Loh P-R, Palamara PF, Price AL. Fast and accurate long-range phasing in a UK Biobank cohort. Nature Genetics. 2016;48(7):811–6.

25. Bycroft C, Freeman C, Petkova D, Band G, Elliott LT, Sharp K, et al. The UK Biobank resource with deep phenotyping and genomic data. Nature. 2018;562(7726):203-9.

26. Shin DM, Hwang MY, Kim BJ, Ryu KH, Kim YJ. GEN2VCF: a converter for human genome imputation output format to VCF format. Genes Genomics. 2020;42(10):1163–8.

27. Ge T, Chen C-Y, Ni Y, Feng Y-CA, Smoller JW. Polygenic prediction via Bayesian regression and continuous shrinkage priors. Nature Communications. 2019;10(1):1776.

28. Park CY, Jo G, Lee J, Singh GM, Lee JT, Shin MJ. Association between dietary sodium intake and disease burden and mortality in Koreans between 1998 and 2016: The Korea National Health and Nutrition Examination Survey. Nutr Res Pract. 2020;14(5):501–18.

29. Park HK, Lee Y, Kang BW, Kwon KI, Kim JW, Kwon OS, et al. Progress on sodium reduction in South Korea. BMJ Glob Health. 2020;5(5).

30. Willer CJ, Li Y, Abecasis GR. METAL: fast and efficient meta-analysis of genomewide association scans. Bioinformatics. 2010;26(17):2190–1.

31. Terry M. Therneau PMG. Modeling survival Data : Extending the Cox Model: Springer; 2000.

